# Implementation of acute flaccid paralysis surveillance for polio eradication in Ethiopia: a qualitative study

**DOI:** 10.64898/2026.04.27.26351901

**Authors:** Wakgari Deressa, Selamawit Hirpa, Anna Kalbarczyk, Svea Closser, Assefa Seme, Olakunle Alonge

## Abstract

Ethiopia has implemented acute flaccid paralysis (AFP) surveillance for nearly three decades as a core polio eradication strategy, yet remains at risk of outbreaks, particularly in pastoralist and conflict-affected areas. As Global Polio Eradication Initiative support declines, understanding factors affecting AFP surveillance and sustainability is critical. This study assessed facilitators, barriers, and adaptive strategies influencing AFP surveillance implementation in Ethiopia. A qualitative study using semi-structured interviews was conducted with 43 participants who had been involved in AFP surveillance for at least 12 months between 1996 and 2018. Guided by the Consolidated Framework for Implementation Research, participants were purposively chosen from the Ministry of Health, regional health bureaus, zonal and district health offices, and included surveillance officers, program managers, and frontline health workers from governmental and partner organizations. Data were analyzed thematically using deductive and inductive approaches in NVivo version 12. AFP surveillance implementation in Ethiopia was influenced by multilevel facilitators and barriers. Strong leadership, organizational structures, and partnerships with global and community actors supported coordination and resource mobilization. Community-based networks, including local volunteers and motivated health workers enhanced case detection and reporting in hard-to-reach areas. However, performance was constrained by high staff turnover, logistical challenges, limited subnational resources, weak supervision, and socio-cultural. Geographic inaccessibility and insecurity further limited implementation. Frontline health workers and volunteers used various adaptive strategies such as community engagement, informal reporting, and context-specific logistical solutions, to sustain surveillance activities. Continued reliance on external support posed a concern for long-term sustainability. Strong organizational systems and community engagement can sustain AFP surveillance in resource-limited settings. However, declining external support is a concern for sustainability. Integrating AFP surveillance into broader health systems, increasing domestic investment, and strengthening community-based approaches are essential for long-term resilience.

## Introduction

Surveillance of acute flaccid paralysis (AFP) is a cornerstone of the Global Polio Eradication Initiative (GPEI), launched by the World Health Assembly in 1988 to eradicate wild poliovirus (WPV). AFP surveillance identifies suspected polio cases among children under 15 years, characterized by the sudden onset of muscle weakness or paralysis, often in the limbs. The process involves laboratory confirmation of these cases through stool samples by accredited laboratories within the Global Polio Laboratory Network [1]. AFP surveillance is implemented through active and passive systems, and is supported by environmental surveillance to improve sensitivity and early outbreak detection [2]. Maintaining sensitive and timely AFP surveillance is essential for detecting poliovirus transmission, monitoring eradication progress, and guiding response, including in setting where WPV has been eliminated but risks of importation and circulating vaccine-derived polioviruses (vVDPVs) persist [2,3].

Despite global progress, important implementation gaps remain. While many countries have achieved their national AFP surveillance targets, substantial subnational disparities persist, particularly in resource-limited and conflict-affected settings [5,6]. In 2023, 71% of priority countries achieved national AFP targets, yet weaknesses in surveillance quality, reporting timeliness, and specimen adequacy were still observed [6]. In Africa, surveillance performance has fluctuated, partly due to COVID-19 pandemic disruptions [5]. Countries in the Horn of Africa, including Ethiopia, Kenya, and Somalia, have faced significant gaps in resources, training, and supervision, which affected AFP surveillance systems [7].

In Ethiopia, AFP surveillance has been central to polio eradication since 1996, contributing to the interruption of indigenous WPV transmission in 2001 [8,9]. National AFP surveillance performance has improved over time, reflecting increased sensitivity and stool specimen adequacy [8]. Recent evidence in some regions showed a highly sensitive AFP surveillance system, with non-polio AFP rates of 3.8–4.8 per 100,000 children under 15, exceeding the stool adequacy rates above 90% [10]. However, surveillance performance remains suboptimal in peripheral and pastoralist regions (Afar, Benishangul-Gumuz, Gambella, and Somali). In these areas, many woredas (districts) fail to meet the 80% stool adequacy threshold, creating ‘silent pockets’ of poliovirus transmission [8]. These weaknesses contribute to continued vulnerability to poliovirus importations and cVDPV outbreaks. With recent reports of cVDPV2 cases, Ethiopia remains classified by the GPEI as an outbreak-prone country [11–14].

Ethiopia’s AFP surveillance operates within the Integrated Disease Surveillance and Response (IDSR) framework, but retains parallel structures with dedicated resources, including staff, funding, and reporting mechanism. As global polio funding declines, there is an increasing need to understand how such systems can be sustained and integrated into broader national surveillance platforms [3,15]. Understanding these implementation dynamics is critical for improving surveillance performance. Existing studies have largely focused on quantitative performance indicators, providing limited insight into the contextual, organizational, and behavioral factors that influence implementation [7,8,16]. However, there is limited qualitative and theory-informed evidence on the implementation of AFP surveillance in Ethiopia. Consequently, there is a critical gap in understanding frontline actors and stakeholders experience, adapt, and respond to challenges within the Ethiopian health system.

To address this gap, this study applied the Consolidated Framework for Implementation Research (CFIR) to examine the multilevel barriers, facilitators, and adaptive strategies influencing AFP surveillance implementation in Ethiopia [17]. CFIR provides a comprehensive framework for systematically assessing how intervention characteristics, inner and outer settings, individual actors, and implementation processes interact to shape program implementation. The findings are expected to generate evidence to strengthen AFP surveillance and support its sustainable integration into broader surveillance systems for polio and other vaccine-preventable diseases (VPDs).

## Methods

### Study setting

This study is part of the ‘Synthesis and Translation of Research and Innovations from Polio Eradication’ (STRIPE) project [9,18]. The study was conducted in Ethiopia, focusing on Addis Ababa, Oromia, Somali, Gambella, and Benishangul-Gumuz (BG) regions. These areas have diverse demographic, geographic, socio-economic, and health systems, and include cross-border areas and places at risk of polio outbreaks due to cVDPVs and imported WPV.

In Ethiopia, AFP surveillance is organized from the national to the community level to ensure rapid detection of WPV and cVDPV2. Nationally, the Ministry of Health (MoH) provides policy oversight, while the Public Health Emergency Management (PHEM) unit at the Ethiopian Public Health Institute (EPHI) is responsible for data management and laboratory confirmation through a WHO-accredited national polio laboratory, with technical support from the WHO and other GPEI partners. At regional and zonal levels, surveillance officers coordinate implementation, with particular emphasis on high-risk regions such as Somali and border areas. At the district (woreda) level, health offices work directly with health facilities to find, report, and investigate AFP cases.

Ethiopia’s Health Extension Program (HEP) provides the backbone for community-based surveillance at the frontline [19]. Healthcare providers and Health Extension Workers (HEWs) identify AFP symptoms, initiate reporting, and facilitate stool sample collection. At the community level, HEWs extend surveillance through community volunteers such as Women Development Armies (WDAs) to the household level. Religious and community leaders support AFP case detection and reporting. The network of CORE Group Partners Project (CGPP) helps with active surveillance in remote or hard-to-reach areas [16].

### Study design

This study employed a qualitative implementation research design using semi-structured interviews with both open- and closed-ended questions. The study was guided by the CFIR to systematically explore the multi-level dynamics of AFP surveillance [17]. This approach allowed for a comprehensive assessment of organizational readiness, internal and external facilitators and barriers, and the adaptive strategies used by actors across the health system hierarchy. This study was reported in accordance with the Standards for Reporting Qualitative Research (SRQR) and the completed checklist is provided in S1 Checklist.

### Study participants, sampling and recruitment

Survey respondents were adults (≥18 years) with at least 12 months of experience in AFP surveillance between 1996 and 2018. A purposive sampling approach designed for complex and multi-institutional partnerships like GPEI was used to identify survey respondents [9,20]. Respondents were selected to ensure representation from government and non-governmental organizations (NGOs) at national, regional, zonal, and woreda levels, including managers, surveillance officers, and frontline health workers, capturing contextual and socioecological variation in AFP surveillance implementation.

### Data collection

A semi-structured questionnaire developed in English was used to collect data. It covered five major internal domains: individual, organizational, polio program, implementation process characteristics, and five external domains: social, economic, political, technological, and environmental factors. Open-ended questions allowed respondents to share experiences across CFIR domains, which were analyzed as internal and external facilitators and barriers to AFP surveillance. The questionnaire also collected information on strategies used to address facilitators and barriers, emphasizing adaptive strategies and best practices.

The questionnaire underwent a series of revisions, translated into Amharic, and back-translated into English to ensure accuracy and consistency. Six interviewers with postgraduate degrees and qualitative research experience were recruited and received three days training. SH participated in the data collection. The training was given by the research team, who had an extensive experience in both qualitative and quantitative studies. Before fieldwork, the questionnaire was piloted, and minor changes were made for clarity and practicality. Interviewers identified and contacted eligible participants, explained the aim of the study, scheduled interviews, and met respondents in person on the interview day. All interviews were conducted face-to-face in Amharic between December 10, 2018 and January 15, 2019. Interviews lasted 60-90 minutes, after which interviewers summarized key points and prepared field notes for team discussion to validate the data before the next interview.

To ensure trustworthiness and rigor, member checking was conducted during data collection by summarizing key responses at the end of each interview and confirming their accuracy and interpretation with participants. Peer debriefing was also undertaken within the research team to review emerging themes and interpretations. These strategies were employed to strengthen the credibility and consistency of the analysis.

### Data analysis

Although data were collected on all polio eradication program goals, only AFP surveillance data were analyzed in this paper. Interviews were transcribed verbatim in Amharic and translated into English for analysis. Data were coded across four CFIR inner setting domains (implementation process, organizational characteristics, polio program, and individual characteristics) and five outer setting domains (social, political, economic, technological, and geographic or environmental factors). Data were managed using NVivo version 12 and analyzed through a hybrid approach of deductive and inductive thematic coding. Deductive coding was guided by CFIR, while inductive coding captured emergent themes such as unintended consequences and cross-program learning.

To ensure rigor, we employed triangulation of perspectives across different administrative levels and peer debriefing among research team to reach a consensus on the final thematic map. The findings are presented by first outlining the facilitators, followed by the barriers and the corresponding adaptive strategies employed to address them along the primary internal and external domains. Selected verbatim quotes from the interview data are presented to illustrate the themes.

### Ethics statement

The study protocol for the project titled ‘‘Implementation Science for Global Health: Lessons Learned from the Global Polio Eradication Initiative in Ethiopia” was approved by the Institutional Review Board (IRB) of the College of Health Sciences at Addis Ababa University (Protocol No: 081/18/SPH; dated December 04, 2018;). The study protocol was also approved by the IRB at the Johns Hopkins Bloomberg School of Public Health (IRB No: 00008721; dated May 14, 2018;).

Consent was obtained from all respondents prior to interviews. Each participant provided written informed consent and permission for interview. Any identifying personal information from the participants was not obtained. Confidentiality was maintained by de-identifying transcripts and securing data storage.

## Results

### Characteristics of respondents

Of the 101 survey respondents, 43 (42.6%) had worked in AFP surveillance at various levels of the health system between 1996 and 2018 (Table 1). About 88% were men, with an average of 9.5 years of experience. The majority worked at zonal (67.4%), or woreda (41.9%) level, and 67.4% were from government institutions. Almost all (93%) had worked in Ethiopia as supervisors, surveillance officers, program managers, or frontline health workers.

**Table 1.** Characteristics of respondents.

| Variable (n=43) | Number of respondents (%) |
| --- | --- |
| **Gender** |  |
| Male | 38 (88.4) |
| Female | 5 (11.6) |
| **Years served in polio program** |  |
| <5 years | 4 (9.3) |
| 5-10 years | 28 (65.1) |
| 11-15 years | 4 (9.3) |
| >15 years | 7 (16.3) |
| **Levels worked in polio program\*** |  |
| Global or national | 6 (13.9) |
| Regional | 12 (27.9) |
| Zonal | 29 (67.4) |
| Woreda (or District) | 18 (41.9) |
| Health facility | 10 (23.3) |
| **Organizational representation\*** |  |
| Government | 29 (67.4) |
| GPEI core partner | 14 (32.6) |
| NGO implementing partner | 9 (20.9) |
| **Countries worked** |  |
| Ethiopia | 40 (93.0) |
| Other | 3 (7.0) |

| **Primary role in polio program\*** |  |
| --- | --- |
| Supervisor | 16 (37.2) |
| Surveillance officer | 14 (32.6) |
| Frontline health worker | 13 (30.2) |
| EPI manager | 12 (27.9) |
| Program manager | 8 (18.6) |
| Program officer | 8 (18.6) |
| **Position at the time of survey** |  |
| Program manager/officer | 20 (46.5) |
| EPI manager | 11 (25.6) |
| Frontline health worker | 6 (14.0) |
| Other | 6 (14.0) |

*\*Many respondents served multiple roles in polio eradication activities at different times*

### Domains of key internal facilitators and barriers

Internal factors refer to characteristics within the health system itself, including individuals, organizations, and the processes used to implement AFP surveillance. Table 2 summarizes the key internal facilitators and barriers to AFP surveillance with definitions across system levels. The top internal facilitator reported was the implementation process, followed by organizational and polio program-related facilitators. In contrast, individual-level factors were the most commonly cited internal barriers, while fewer respondents identified barriers related organizational structures or implementation processes. Illustrative examples of internal facilitators and barriers, organized by CFIR domains and constructs, are provided in the supplementary materials (S2 and S3 Tables).

**Table 2.** Key internal facilitators and barriers to AFP surveillance (n=43)

| **Facilitators\*** | **Definitions** | **Number of respondents (%)** |
| --- | --- | --- |
| Implementation process | Well-planned, executed, monitored, and adapted activities facilitated efficient AFP surveillance activities | 16 (37.2) |
| Organizational | Supportive structures, leadership, resources, and coordination within the organization facilitated effective AFP case detection and reporting | 11 (25.6) |
| Polio program | Adoption of surveillance tools, guidelines, and technologies that facilitated case detection, reporting, and monitoring | 11 (25.6) |
| Individual | Positive attitudes, adequate knowledge, relevant training, and skills of health workers and organizational staff facilitated timely identification, reporting, and investigation of AFP cases | 3 (7.0) |
| **Barriers\*** |  |  |
| Individual | Attitudes, knowledge, training, or skills that hinder health workers' and organizations' ability to implement AFP surveillance effectively | 10 (23.3) |
| Polio program | Inadequate, inappropriate, or poorly adopted program activities, strategies, or technologies that limit the effectiveness of AFP surveillance | 7 (16.3) |
| Implementation process | Gaps or inefficiencies in planning, execution, monitoring, reflection, or adaptation of activities that impede the effective implementation of AFP surveillance | 4 (9.3) |
| Organizational | Structural, resource, or procedural limitations within the organization that obstruct effective AFP surveillance | 1 (2.3) |

\*Respondents could select few or multiple domains as facilitators or barriers to AFP surveillance; therefore, percentages may not be 100%.

#### Implementation process

AFP surveillance implementation was supported by a structured and proactive operational approach that included prioritization of health facilities based on patient volume and routine verification of facility registers. This enabled surveillance teams to move beyond passive reporting toward active case finding and zero-reporting verification. As one district surveillance officer explained:

> *“We classify and prioritize health centers based on patient volume. We schedule visits accordingly and review their registration books to check for cases that have signs of polio. If a case was missed, we contact the family using the address and take stool samples.”* (District Surveillance Officer, Addis Ababa).

However, implementation was undermined by frequent turnover of trained health workers, which disrupted continuity in surveillance activities and weakened reporting consistency. As noted by an implementing partner:

> *“We provide training on surveillance to health workers. But these people leave the area after sometime, that makes our work challenging.”* (AMREF NGO Partner, National).

To address this challenge, the program adapted by shifting part of the case detection responsibility to community-level structures. Community volunteers function as early warning reporters, particularly in remote and hard-to-reach areas where formal health workers coverage is limited, thereby sustaining surveillance continuity. A district manager described this approach:

> *“In each village, there is a designated community volunteer responsible for reporting notifiable diseases in hard-to-reach areas. If any such disease occurs, she/he reports to the woreda so that timely action can be taken.”* (District EPI Program Manager, BG).

#### Organizational characteristics

The organizational strength of AFP surveillance was anchored in the formal integration of GPEI structure within the national PHEM structure. This created a clear hierarchical reporting system from the national level to the kebele (sub-district) level, and supported multi-sectoral coordination in surveillance implementation, as described by a zonal officer.

> *“A strong structure from the MoH to the lowest level sub-district (kebele) established and working on the effective implementation of the program. The close collaboration of different organizations together with the governmental organization in polio eradication creates a favorable condition for surveillance activity*.” (Zonal Surveillance Officer, Gambella).

Despite this, implementation was constrained by severe human resource and logistical limitations at subnational levels. A single PHEM officer often covered large geographic areas with numerous kebeles, faced limited communication resources, and worked without adequate operational support. These challenges were compounded by the absence of regional laboratory capacity, which delayed specimen transport and diagnosis. A zonal officer noted:

> *“PHEM is a very broad area but only one PHEM officer is assigned to take all responsibilities at zonal and woreda level. Some woredas [district] have up to 74 kebeles [sub-district]. The officer at woreda has to call every HEW to communicate on reports and fill mobile airtime at his/her own expense.”* (Zonal Surveillance Officer, BG).
>
> *“We also do not have a regional laboratory which work on the diagnosis of the polio virus. Because of poor transportation from rural areas to the town, and then to the national laboratory, it takes more days for the stool to be diagnosed.”* (Frontline Health Worker, BG).

To address these constraints, the program relied on continuous recruitment and training of surveillance personnel and provided targeted financial and logistical support to woreda surveillance officers to strengthen community-based surveillance activities.

> *“Since human power for surveillance is a key requirement, we do our best in identifying resources to recruit manpower. We continuously conduct training for health workers on surveillance and provide financial support to woreda surveillance officers so they can conduct community surveillance.”* (WHO Surveillance Coordinator, BG).

#### Polio program characteristics

The prioritization of polio eradication at the national level ensured sustained political attention, resource allocation, and coordinated action among implementing partners, creating a supportive environment for AFP surveillance implementation. As noted by a partner organization:

> *“The collaboration of different organizations together with the governmental organization in polio eradication creates a favorable condition for surveillance activity.”* (CGPP Partner, Gambella).

However, the specialized nature of AFP surveillance limited ownership at lower levels, as responsibility was often concentrated among a small group of designated personnel. This narrow engagement limited broader community participation and weakened local-level follow-up mechanisms. A frontline health worker explained:

> *“During the past few years, the polio surveillance only came to a few responsible people, and this has created the community not to be involved in the eradication of the virus. This had also decreased tracking and follow-up for the surveillance work.”* (Frontline health worker, BG).

To address this, the program strengthened supportive supervision through periodic deployment of experts from the MoH and WHO to mentor local staff and reinforce implementation capacity. Efforts were also made to decentralize technical expertise to improve local detection and response capacity.

> *"Through the coordination of MoH and partners [WHO], we identify resources and provide training and supervision for newly recruited health workers."* (WHO Surveillance Coordinator, BG)

#### Individual characteristics

At the individual level, AFP surveillance performance was facilitated by the perceived value of training and the motivation generated through per-diem incentives. These factors encouraged health workers to actively engage in surveillance activities, particularly regarding the challenging task of sample collection and transport. A frontline health worker noted:

> *“Once the suspected cases are identified, samples will be collected and taken to EPHI [national laboratory]. Health workers that took the sample to the laboratory get incentives that motivate the staff to identify cases.”* (Frontline Health Worker, Addis Ababa).

However, a key behavioral barrier was the prioritization of vaccination activities over surveillance responsibilities. Many health workers often perceived vaccination campaigns as the primary task, resulting in limited engagement in active case detection and reliance on routine reporting rather than active surveillance, as described by respondents:

> *“Health workers conducting the vaccination campaign only focus on the vaccine delivery and the quality of the vaccine, rather than conducting the surveillance using the checklist provided. They just report zero without any consideration.”* (Zonal EPI Coordinator, BG). *“Health workers at health facilities are sometimes reluctant to go to the community and conduct community-based surveillance. Rather, they wait for the community to come to the health facility.”* (CGPP Implementing Partner, BG).

To address these challenges, the program used regular review meetings and on-site mentoring. Supervisors reviewed facility registers with health workers to identify missed cases and provide immediate corrective feedback, turning surveillance gaps into learning opportunities, as noted by regional officers.

> *“We conduct review meeting, refresher trainings and also motivate those health workers with high performance in AFP surveillance.”* (WHO Surveillance Officer, BG).
>
> *“When we go to the health facility for monitoring, we check the registration book for any sign of AFP which is not reported. The health workers accept our comments and are willing to correct their mistake.”* (Regional Cold Chain Officer, BG).

### Domains of key external facilitators and barriers

External factors refer to influences outside the health system that shape AFP surveillance, including social, political, economic, technological, and geographic conditions. Table 3 summarizes the key external facilitators and barriers to AFP surveillance with definitions across system levels. Among these facilitators, social factors, including community awareness and cooperation, were most frequently reported, while geographic and environmental factors, such as remoteness and difficult terrain, followed by technological and political factors, were the most commonly cited external barriers. Illustrative examples, organized by CFIR domains and constructs, are provided in the supplementary material (S4 Table).

**Table 3.** Key external facilitators and barriers to AFP surveillance (n=43)

| Facilitators* | Definitions | Number of respondents (%) |
| --- | --- | --- |
| Social | Community awareness, norms, beliefs, cooperation, network, and acceptance that facilitated reporting of AFP cases and community-based surveillance | 19 (41.2) |
| Political | Supportive political climate and leadership at all levels that facilitated coordination and prioritization of AFP surveillance | 10 (23.3) |
| Economic | Adequate funding and resource mobilization that facilitated implementation of AFP surveillance activities | 8 (18.6) |
| Technological | Availability of external infrastructure or technological tools that facilitated effective AFP surveillance, such as mobile phones | 2 (4.7) |
| Geographic/<br>environmental | Accessible locations and favorable geographic conditions that facilitated case finding and investigation | 2 (4.7) |
| **Barriers\*** |  |  |
| Geographic/<br>environmental | Physical inaccessibility, challenging terrain, remoteness, or environmental conditions that impede effective AFP surveillance | 14 (32.6) |
| Technological | Weak external infrastructure, poor communication systems, or delayed laboratory feedback that hindered AFP surveillance | 13 (30.2) |
| Political | Insecurity, weak political support, or instability that challenged implementation of AFP surveillance | 12 (27.9) |
| Economic | Insufficient funding, limited revenue sources, or lack of financial support constrained AFP surveillance activities | 10 (23.3) |
| Social | Prevailing social norms, beliefs, and misconceptions in the community that limit AFP surveillance activities | 9 (20.9) |

\*Respondents could select few or multiple domains as facilitators or barriers to AFP surveillance; therefore, percentages may not be 100%.

#### Social environment

The broader social environment facilitated AFP surveillance through strong community engagement and social capital. Collaboration with community members, including religious and clan leaders supported case identification and reporting, particularly in pastoralist settings, as noted by a CGPP partner:

> *"People are cooperative once they understand the purpose and accept your activity, especially if you involve religious and clan leaders. The society has a good communication culture and shares every piece of information about new places they plan to go along with their livestock. We have used this social capital to identify people with AFP symptoms; they could even tell you about such cases living 50 or more kilometers away."* (CGPP Partner, Somali).

Community-based surveillance was further strengthened through the WDAs network, which ensured household-level detection and reporting through established linkages with the formal health system. Information identified at the household level was first reported to WDAs, who then verified and relayed it to health facilities and district (woreda) health offices as needed.

> *"Orientation was provided to WDAs and their networks on the signs and symptoms of AFP*. *When they suspect cases in the community, they report to HEWs, and HEWs to health workers at health centers. The health workers visit the family and collect samples and take it to the central laboratory for investigation."* (HEWs Supervisor, Addis Ababa)

Despite these facilitators, socio-cultural beliefs and norms posed significant barriers to timely case reporting. In some communities, AFP was attributed to spiritual causes, leading families to seek care from traditional or religious leaders outside the formal surveillance system. Furthermore, social norms around confidentiality discouraged reporting of suspected cases within communities, as noted by zonal officer.

> *“Some community members believe AFP is a curse from the creator resulting from sin, and they seek holy water or traditional healers, such as prayers, as a form of treatment.”* (Zonal Health Officer, Oromia).
>
> *“They consider reporting a neighbors’ problem (e.g., a child with paralysis) as exposing confidential matters to non-relatives, and they consider it as a taboo.”* (CGPP Partner, Somali).

To overcome these barriers, the program employed community-based advocacy strategies by engaging religious and clan leaders to improve awareness, counter misconceptions, and promote timely reporting of suspected AFP cases.

> *“We also work with the religious and tribal leaders to teach the community about the polio virus.”* (CGPP Partner, Somali).

#### Political environment

The recognition of polio eradication as a high-level national and international priority created a supportive political climate that ensured the continuous mobilization of resources and technical support for AFP surveillance. As a national PHEM officer noted:

> *"There is a support from national and international partners, and it also gets emphasis from political leadership; resources and technological support have enabled the program to achieve its surveillance goals."* (PHEM Officer, National).

However, insecurity and regional conflicts posed barriers to implementation by restricting physical access to high-risk areas and creating ‘silent zones’ where surveillance activities could not be conducted. A WHO surveillance officer described the operational impact of conflict as follows:

> *“Due to the conflict in some parts of our region, we could not conduct AFP surveillance in certain areas for over two months. In places with security issues, we send people living in that community to conduct surveillance and report to us.”* (WHO Surveillance Officer, BG).

Similarly, a frontline health worker emphasized the broader implication of insecurity on AFP surveillance efforts:

> *"To eradicate polio, security issue is our major problem. Polio is usually found at places where there are security issues. This has inhibited our interventions in the surveillance of AFP cases."* (Frontline Health Worker, BG).

To maintain surveillance in these settings, the program adopted context-specific security strategies, including the use of local community members for reporting and conducting field activities with security escorts when access was possible. A regional surveillance officer explained:

> *“In some areas, we also took security personnel with us and travel to areas where there is a conflict.”* (Regional Surveillance Officer, BG).

#### Economic environment

External funding from global partners supported key surveillance functions, including field operations, diagnostics, and incentives. A frontline health worker noted:

> *“The economic resource supported from WHO, UNICEF and others working on polio eradication helped us to achieve our surveillance goals.”* (Frontline health worker, Oromia).

Despite this external support, insufficient local funding and limited resource allocation at subnational level constrained surveillance implementation, particularly for outreach activities in hard-to-reach areas. An NGO manager highlighted:

> *“Cross-border instability, insufficient budget allocation, high personnel turnover, and inaccessibility of implementation areas are major challenges.”* (NGO Program Manager, National)

To cope with these constraints, frontline actors often relied on informal practices, including the use of personal resources to maintain reporting. For example, community-level actors used their personal mobile phones to report surveillance data from remote areas, as described a district manager:

> *"Those kebeles who do not send the report due to distance sends it by a mobile message (text message) at their own expense."* (District EPI Program Manager, BG).

#### Technological environment

Simple communication technologies facilitated AFP surveillance implementation. Mobile phones, text messaging, and radio communication to report suspected cases, improving timeliness and coordination. In some areas, digital tools such as Open Data Kit (ODK) were introduced to support real-time reporting. However, weak telecommunication networks and limited access to mobile devices constrained timely reporting in some areas. A partner at the national level noted:

> *“Reporting polio cases through phone and the staff took the initiative to use ODK software which is useful to report cases.”* (CGPP Partner, National).

#### Geographic/environmental factors

Geographic and environmental conditions posed significant barriers to AFP surveillance implementation. Respondents described challenges related to hard-to-reach areas, scattered settlements, seasonal flooding, and the mobility of pastoralist populations, all of which limited routine homes visits and case follow-up. To overcome these constraints, surveillance teams adapted context-specific logistical strategies, including the use of locally available transport and deployment of physically capable personnel to access difficult terrain. A regional officer explained:

> *“For hard-to-reach areas, we choose people who can endure harsh environment based on their will. We used traditional transport system such as boat for river, and donkeys for long distances.”* (Regional Surveillance Officer, BG).

## Discussion

The findings show that AFP surveillance is supported by strong organizational structure, sustained political commitment, and active community engagement. However, these strengths are offset by persistent challenges including, workforce turnover, subnational resource constraints, socio-cultural barriers, geographic inaccessibility, and insecurity in conflict-affected areas. These contextual and system-level constraints contribute to variations in surveillance performance, particularly in hard-to-reach and underserved areas. Frontline workers employed a range of adaptive strategies to address these challenges through context-specific strategies, such as community-based surveillance, informal communication mechanisms, and locally appropriate logistical solutions, to sustain surveillance functions. The findings highlight that AFP surveillance performance is driven not only by technical capacity, but by how health systems and communities interact and adapt within complex and resource-constrained environments. These findings also highlight the critical role of adaptive implementation, where frontline workers continuously modify strategies to sustain surveillance under constrained and dynamic conditions.

The findings highlight that Ethiopia’s well-structured AFP surveillance system enabled coordination across national, regional, and community levels, reflecting key CFIR inner setting constructs such as organizational structure and leadership engagement. Strong national leadership and the integration of polio surveillance into the broader PHEM system elevated program priority and responsiveness, consistent with findings from other international settings [21–23]. In addition, partnerships with GPEI core partners, NGOs, and civil society organizations facilitated resource mobilization and strengthened linkages between surveillance and vaccination activities [18,24]. Initiatives such as the CGPP further demonstrate how coordinated partnerships can extend surveillance reach to marginalized and hard-to-reach populations [24,25]. However, while these structural and partnership-driven strengths contributed to national-level performance, they did not fully address persistent subnational gaps, suggesting that strong system design alone is insufficient without adequate local capacity and contextual adaptation. This underscores the importance of translating high-level coordination into effective implementation at lower levels.

However, a critical implementation gap remains between national policy and local reality. While high-level partnerships mobilized substantial resources [24], these vertical investments often failed to address fundamental system constraints, including limited transport, inadequate fuel, and lack of regional laboratory capacity. This reflects CFIR inner setting challenges related to available resources and implementation capacity. Similar patterns have been reported in Nigeria and Pakistan, where polio-specific logistics did not sufficiently strengthen broader health system [22]. The persistent delays in sample transport identified in this study suggest that without domestic investment in core logistics, surveillance systems remain dependent on external donor funding that are declining during the transition phase. This raises important concerns regarding the long-term sustainability of AFP surveillance, underscoring the need for increased domestic investment and stronger integration into routine health systems. Furthermore, gaps in local-level knowledge and inconsistent supervision indicate weaknesses in implementation processes, contributing to underreporting and limiting the effectiveness of otherwise well-designed surveillance systems [26,27]. Collectively, these multilevel constraints help explain the persistent subnational disparities in AFP surveillance performance observed in Ethiopia, including delays in case detection and underreporting in hard-to-reach and conflict-affected areas.

Consistent with global evidence, this study found that health worker motivation is a key determinant of surveillance performance [28]. However, gaps in training, retention, and resource support remain significant barriers, particularly in pastoral and remote areas. Health workers in these settings faced competing demands, often conducting active case searches without dedicated transport budgets or the financial incentives associated with AFP surveillance tasks [29,30]. This reflects CFIR constructs related to individual motivation and competing priorities, and helps explain the observed preference for vaccination campaigns over routine surveillance activities. Addressing these challenges requires strategies to improve retention and job satisfaction through fair incentives, recognition, and supportive supervision [31].

CBS structures, particularly the WDAs, played a critical role in sustaining AFP surveillance at the household level. These frontline volunteers supported active case finding, reporting, and community awareness raising, reflecting CFIR process constructs related to engagement and implementation at the frontline. Their close integration within communities enabled early identification of suspected cases, particularly in areas with limited access to health facilities. This finding is consistent with previous studies in Ethiopia showing that frontline workers are vital for AFP surveillance [27,32]. Globally, CBS has improved AFP case detection through such engagement in countries like Nigeria, Somalia, South Sudan, and India, including a doubling of AFP reporting in some Ethiopian settings [16,33,34]. However, the effectiveness of such approaches depends on sustained support, supervision, and integration within the formal health system to ensure data quality and continuity.

The vertically structured nature of AFP surveillance may limit ownership at lower levels, reflecting CFIR constructs related to intervention complexity and design. This creates ongoing tension between vertical program delivery and broader health system integration, with implications for efficiency and long-term sustainability. While CBS has strengthened local engagement, inconsistent practices and limited feedback mechanisms continue to affect performance in areas with weak supervision [17,32,34]. Addressing these challenges requires strengthening volunteer capacity through improved integration into the formal health system and the provisions of more sustainable incentive mechanisms, rather than reliance on ad-hoc payments, to ensure reliable surveillance in high-risk populations [35].

A key contribution of this study is the finding that social capital functioned as a core component of surveillance implementation with limited availability of health facilities. In pastoralist regions of Ethiopia, clan leaders and religious figures were not only supportive actors but played a central role in enabling case detection and information flow, reflecting the CFIR outer setting constructs related to community networks and social context. In these contexts, trust and established communication channels were as critical as health system inputs [33]. Unlike settings where community volunteers act primarily as passive reporters, Ethiopia’s approach positioned traditional and religious leaders as active intermediaries who legitimized surveillance activities and facilitated reporting across dispersed and mobile populations. This suggests that leveraging existing social structures can enhance surveillance reach in hard-to-reach and insecurity areas, where health facility-based approaches are insufficient. Such contextually embedded strategies may offer a transferrable model for similar settings, although their effectiveness depends on alignment with local social norms and power structures [35].

Geographic and environmental barriers, including difficult terrain, seasonal flooding, and dispersed populations, continue to limit access to facilities and constrain community outreach activities. In addition, political instability in some regions has disrupted surveillance operations, reinforcing CFIR outer setting challenges related to context and external shocks. Despite these constraints, CBS networks were able to sustain case detection and reporting when formal facility-based systems were compromised, consistent with findings from other conflict-affected settings [27,36,37]. These findings align with emerging health system resilience frameworks, which emphasize the role of community-based structures in maintaining essential functions during crises [38,39]. Notably, while AFP surveillance was disrupted during the COVID-19 pandemic across East and Southern Africa [40,41], the CBS mechanisms identified in this study appear to have provided a degree of continuity. In this context, simple technological solutions, such as mobile-based reporting, further strengthened system adaptability by enabling timely communication and case detection in resource-constrained areas [31,42,43].

## Strengths and limitations

A major strength of this study is its application of the CFIR framework to capture over 20 years of institutional memory, moving beyond quantitative indicators to reveal the practical adaptive strategies that sustained surveillance. The use of qualitative methods enabled in-depth exploration of contextual and operational realities that are not reflected in routine surveillance indicators. However, the findings are based on self-reported experiences, which may be subject to recall bias given the 1996–2018 timeline. To mitigate this, we employed data triangulation by cross-referencing participant reflections with official program reports and validating historical themes across multiple administrative levels. The absence of audio recording may have limited the depth and verbatim accuracy of the qualitative data. Additionally, while the specific cultural contexts (e.g., pastoralist clan structures) limits generalizability, the study provides important insights for strengthening surveillance systems in similar low-income settings in transition phase of polio endgame.

## Conclusions

This study demonstrates that the performance of AFP surveillance in Ethiopia is influenced by the interaction of strong organizational design combined with contextual factors, and adaptive responses at multiple levels of the health system. Beyond technical capacity, sustained surveillance depends on how well structures, resources, and community systems align within complex and resource-limited settings. Strengthening the integration of AFP surveillance into routine health services, alongside sustained investment in local capacity and community-based mechanisms, is essential for maintaining sensitivity and responsiveness. These measures are critical for sustaining progress toward polio eradication and for reinforcing broader disease surveillance systems in similar contexts.

## Acknowledgements

We would like to thank the study team members from the Addis Ababa University School of Public Health and the STRIPE Consortium. We also gratefully acknowledge our colleagues at the Johns Hopkins Bloomberg School of Public Health for their valuable inputs. Finally, we are grateful to all data collectors, supervisors, and coordinators who contributed to this study.

## Author contributions

**Conceptualization**: Olakunle Alonge, Anna Kalbarczyk, Svea Closser.

**Data curation:** Wakgari Deressa.

**Formal analysis:** Wakgari Deressa, Selamawit Hirpa.

**Investigation:** Wakgari Deressa, Assefa Seme, Selamawit Hirpa.

**Methodology:** Olakunle Alonge, Wakgari Deressa, Selamawit Hirpa, Anna Kalbarczyk, Svea Closser, Assefa Seme.

**Project administration:** Wakgari Deressa, Assefa Seme, Selamawit Hirpa, Anna Kalbarczyk, Olakunle Alonge.

**Supervision:** Wakgari Deressa, Assefa Seme, Selamawit Hirpa.

**Writing- original draft:** Wakgari Deressa.

**Writing- review and editing:** Olakunle Alonge, Wakgari Deressa, Selamawit Hirpa, Anna Kalbarczyk, Svea Closser, Assefa Seme.

## Data availability statement

Data is provided within the manuscript or supplementary information files. In addition, the full datasets used and analyzed in the current study are available from the corresponding author upon reasonable request.

## Consent for publication

Not applicable

## Supporting information

**S1 Checklist. SRQR Reporting Checklist**

**S2 Table. Internal facilitators to AFP surveillance**

**S3 Table. Internal barriers to AFP surveillance**

**S4 Table. External facilitators and barriers of AFP surveillance**

